# Effects of (Un)lockdown on COVID-19 transmission: A mathematical study of different phases in India

**DOI:** 10.1101/2020.08.19.20177840

**Authors:** R. Kumar, M. Z. Malik, S. R. Shah

## Abstract

The novel coronavirus (SARS-CoV-2), identified in China at the end of the December 2019 is causing a potentially fatal respiratory syndrome (COVID-19), has meanwhile led to outbreak all over the globe. India has now become the third worst hit country globally with 16,38,870 confirmed cases and 35,747 confirmed deaths due to COVID-19 as of 31 July 2020. In this paper we have used mathematical modelling approach to study the effects of lockdowns and un-lockdowns on the pandemic evolution in India. This, study is based on SIDHARTHE model, which is an extension of classical SIR (Susceptible-Infected-Recovered) model. The SIDHARTHE model distinguish between the diagnosed and undiagnosed cases, which is very important because undiagnosed individuals are more likely to spread the virus than diagnosed individuals. We have stratified the lockdowns and un-lockdowns into seven phases and have computed the basic reproduction number R_0_ for each phase. We have calibrated our model results with real data from 20 March 2020 to 31 July 2020. Our results demonstrate that different strategies implemented by GoI, have delayed the peak of pandemic by approximately 100 days. But due to underdiagnosis of the infected asymptomatic subpopulation, a sudden outbreak of cases can be observed in India.

## Introduction

Throughout the human history, infectious deceases have been causing morbidity and mortality. In the last week of December 2019, an infectious disease, causing potential fatal respiratory syndrome, was identified as the novel Coronavirus 2019 (COVID-19) in Wuhan, the capital of Hubei, China. COVID-19 is caused by severe acute respiratory syndrome Coronavirus 2 (SARS-CoV-2) [1–2]. The World Health Organization declared COVID-19 a pandemic on 11 March 2020[3].

India reported its first case of COVID-19 on 30 January 2020, followed by two cases in the month of February, with travel history to COVID-19 affected countries [4]. In March, the transmission grew after several native people and tourists with travel histories to affected countries, and their contacts, tested positive. As the pandemic progressed through the world, Indian government responded with thermal screening of passengers arriving from COVID-19 affected countries. In March, the Indian government issued recommendations regarding social distancing and imposed travel restrictions. On 22^nd^ March, the Government of India (GoI) announced complete lockdown in 82 districts of 22 states of the country where confirmed cases were reported. On 24^th^ March, the Prime Minister of India announced a complete 21-day national lockdown 1 to contain the pandemic. Lockdown 2 was announced on 14^th^ April for 18 days, with conditional relaxations in areas with lower spread from 20^th^ April. On 29^th^ April, the Government issued guidelines for the states to allow inter-state movement of the stranded people. On 1^st^ May, GoI extended lockdown till 17^th^ May, with some more conditional relaxations on the basis of red, orange and green zones. On 17^th^ May, GoI announced another lockdown from 18^th^ to 31^st^ May, with some additional relaxations. The red zones were further divided into containment and buffer zones. GoI issued guidelines on 30^th^ May to reopen the country in various (three) phases from 1June onward. The first phase of reopening was termed as “Unlock 1.0” and permitted shopping malls, religious places, hotels and restaurants to reopen from 8 June. Phase II of Unlock began on 1^st^ July with night curfews. The lockdown measures were only imposed on containment zones [5].

The first human coronavirus outbreak was recorded in 1965 – HCoV-229E, followed by two outbreaks of similar capacity – SARS-CoV and MERS-CoV in 2003 and 2012, respectively [6]. The SARS-CoV-2 is identified as beta-coronavirus belonging to the family of Coronaviridae. Unlike SARS-CoV and MERS-CoV, SARS-CoV-2 shows peculiar epidemiological traits [7]. The COVID-19 transmission depends upon the susceptible population and its infectious agent. Its transmission dynamics can be studied with the help of mathematical models to quantify the possible disease control and mitigation strategies. Several mathematical models have been developed to predict the dynamics of infectious diseases [8–13]. SIR is a widely used deterministic model that describes human to human transmission in three stages: Susceptible(S), Infected(I) and Recovered(R)[14]. Lin et al. exploited SEIR(Susceptible-Exposed-Infected-Recovered) to capture the course of COVID-19 in Wuhan, China [15]. For Covid-19 transmission in India, Mandal et al. developed an extended SEIR model for optimistic and pessimistic scenario [16]. For optimistic scenario the basic reproduction number (R_0_) was 1.5 and in pessimistic scenario, R_0_ was 4. Various mathematical models have been used to predict the COVID-19 pandemic in India [17–21]. Giordano et al. extended classical SIR to SIDARTHE model [22] to predict the course of epidemic in Italy. Their model incorporates the population wide interventions and their effects on infection spread.

In the present study, we generalized the SIDARTHE model to the Indian scenario of COVID-19 that incorporates four phases of lockdown and two phases of un-lockdown. The main novelty of this model lies in including both detected and undetected cases. We have considered both, diagnosed and non-diagnosed cases of COVID-19 in the community. Variability in the transmission between symptomatic and asymptomatic population has been taken into account for every lockdown and un-lockdown period. Basic reproduction number has been computed using next generation matrices. We also estimate the basic reproduction number R_0_ for different phases of lockdown and un-lockdown. Effects of different strategies on the pandemic dynamics have been analyzed by varying the corresponding parameters. Model results have been calibrated with data (from 20 March to 31 July 2020) for cumulative infected cases, cumulative recovered and cumulative deaths.

## Material and Methods

### The data

We have collected the data for diagnosed infected cases, diagnosed recovered and total deaths due to COVID-19 in India for the period of 20 March to 31 July 2020. The real time data for cumulative infected cases and deaths is obtained from WHO website [27]. Data for recovered cases is obtained from relevant sources [28]. Model results have been compared with the real data for total infected cases, recovered cases and total deaths.

### Mathematical formulation of COVID-19 pandemic

We have exploited the SIDARTHE model [22] to see the effects of different lockdowns and un-lockdowns on the number of cases of COVID-19 in India. The total population is subdivided into eight epidemiological classes: S, Susceptible (not infected class); I, Infected (Asymptomatic not detected class); A, Asymptomatic (infected detected class); S_u_, Symptomatic (infected not detected class); D, Detected (infected detected class); C, Critically ill (infected detected class); M, Mortal (detected infected dead class); R, Recovered (detected and undetected infected class). There are very few peculiar cases getting reinfected, but the rate of reinfection is negligible [23], so we haven’t considered the case of recovered people becoming susceptible again. A flowchart of interaction among different classes is shown in **figure 1**.

**Figure 1:**
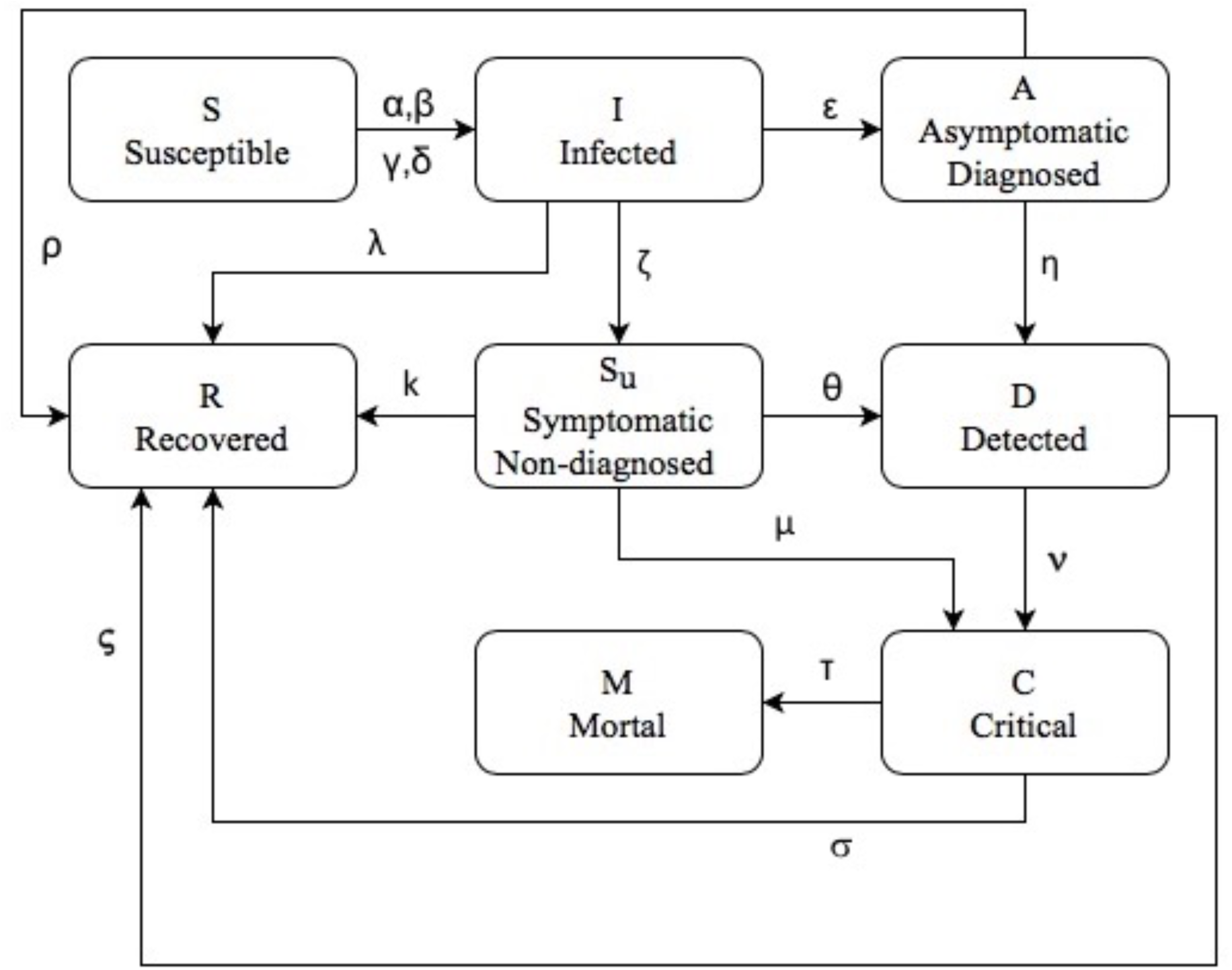
Flowchart of interaction among different class of infection in mathematical model

Time evolution of the population in each class is described by the system of eight ordinary differential equations:

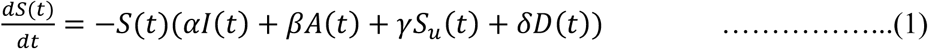

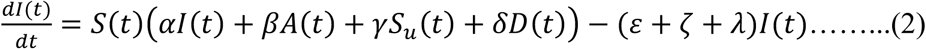

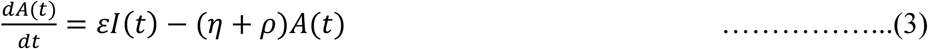

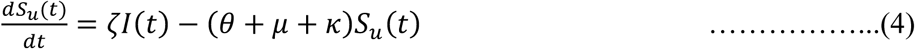

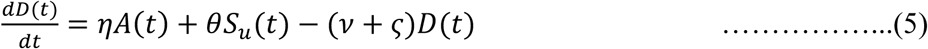

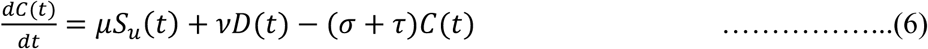

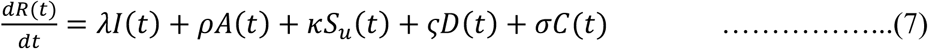

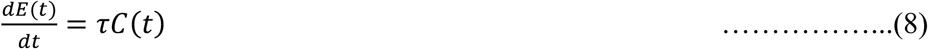

where uppercase letters represent the fraction of population in each class and Greek letters represent the transmission and transition parameters. In particular, the parameters are given in **Table 1**.

**Table 1:**
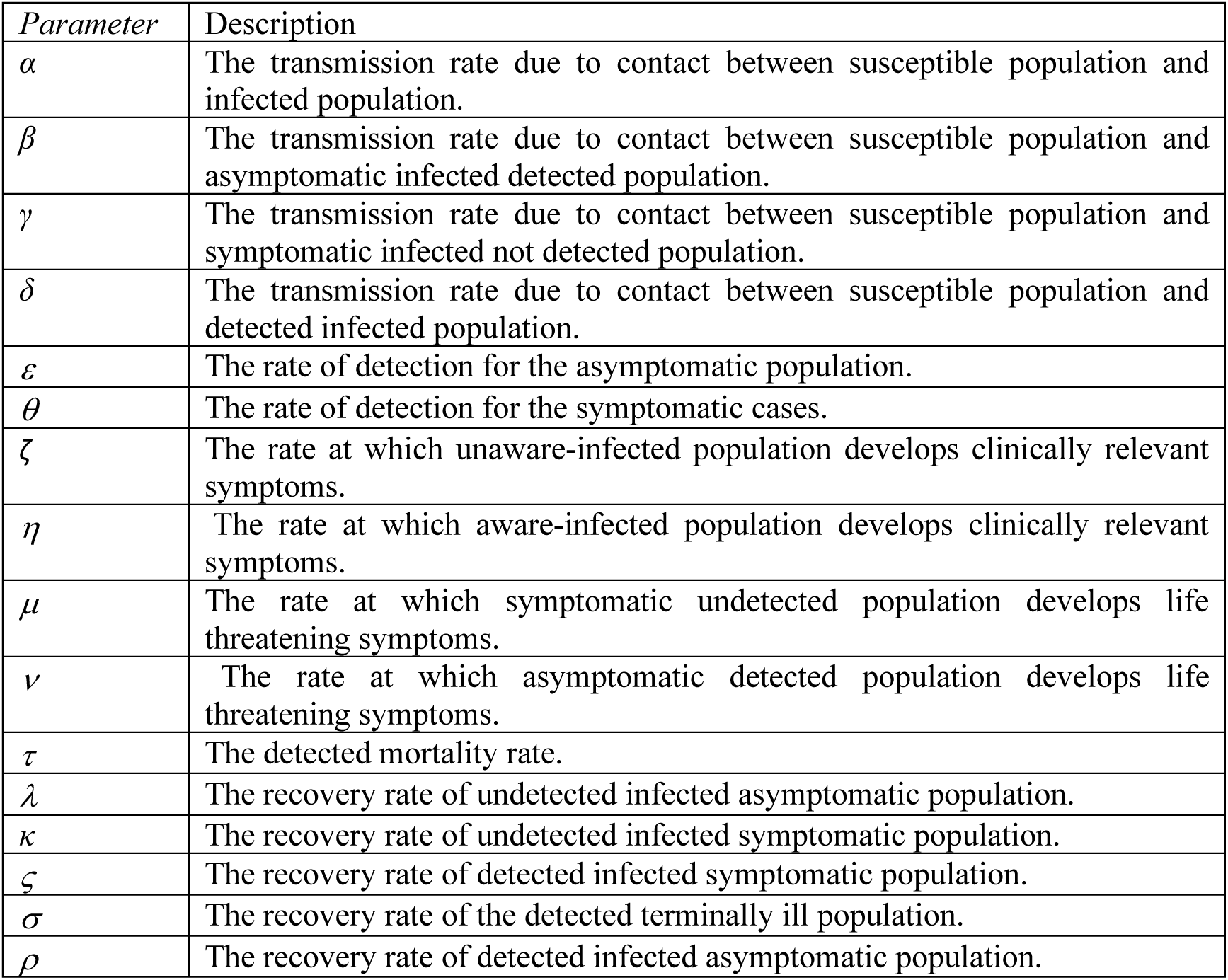
Parameter and descriptions used in our model.

| Parameter | Description |
| --- | --- |
| $\alpha$ | The transmission rate due to contact between susceptible population and infected population. |
| $\beta$ | The transmission rate due to contact between susceptible population and asymptomatic infected detected population. |
| $\gamma$ | The transmission rate due to contact between susceptible population and symptomatic infected not detected population. |
| $\delta$ | The transmission rate due to contact between susceptible population and detected infected population. |
| $\varepsilon$ | The rate of detection for the asymptomatic population. |
| $\theta$ | The rate of detection for the symptomatic cases. |
| $\zeta$ | The rate at which unaware-infected population develops clinically relevant symptoms. |
| $\eta$ | The rate at which aware-infected population develops clinically relevant symptoms. |
| $\mu$ | The rate at which symptomatic undetected population develops life threatening symptoms. |
| $\nu$ | The rate at which asymptomatic detected population develops life threatening symptoms. |
| $\tau$ | The detected mortality rate. |
| $\lambda$ | The recovery rate of undetected infected asymptomatic population. |
| $\kappa$ | The recovery rate of undetected infected symptomatic population. |
| $\varsigma$ | The recovery rate of detected infected symptomatic population. |
| $\sigma$ | The recovery rate of the detected terminally ill population. |
| $\rho$ | The recovery rate of detected infected asymptomatic population. |

### Qualitative analysis of the Mathematical Model

The basic reproduction number, R_0_ is the most important measure which quantifies the disease invasion or extinction in a population [24]. In this section we obtain the basic reproduction number for our model using Next Generation Matrix (NGM). The NGM is the natural basis for the definition and calculation of R_0_ where finitely many different categories of individual are recognised.

### The basic Reproduction number (R_0_)

The basic reproduction number R_0_ is arguably the most significant quantity in infectious disease epidemiology. It is the average number of secondary infections produced by a typical case of infection in a population where everyone is susceptible. R_0_ provides insight when designing control intervention for established infection.

The above system of differential equations (1–8) is compartmental system and has the mass conservation property [25] i.e.,

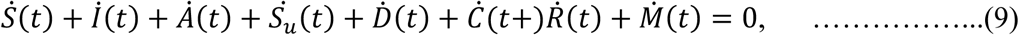

hence, the sum of the states (total population) is constant.

Since the variables denote population fraction, we can assume:

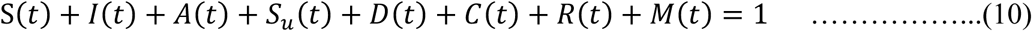

where 1 denotes the total population.

This system (1–8) has five infected states, I(t), A(t), S_u_(t), D(t) and C(t) and three uninfected states, S(t), R(t) and E(t). At the infection free steady state, I(t)= A(t)= S_u_(t)= D(t)= C(t)= 0 with S(t)+ R(t)+E(t) = 1. Hence linearization of equations corresponds to infected states, lead to the linear system

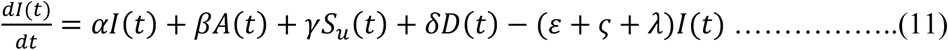

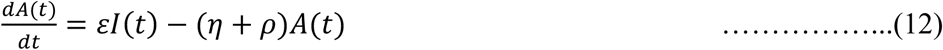

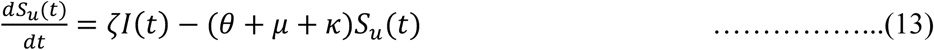

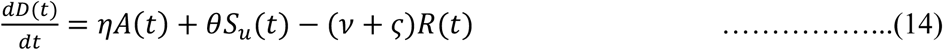

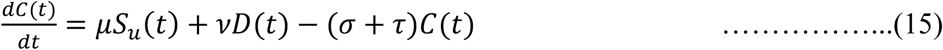

We will refer to this system of ODEs as the linearized infection subsystem.

Let us assume x = (I, A, S_u_, D, C) ’, where prime denotes the transpose, then we can write the linearized infection subsystem in the form

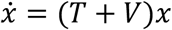

The matrix T corresponds to the transmissions, describing the production of new infections. And matrix V corresponds to the transitions, describing changes in state. The dominant eigenvalue of the matrix K = –TV^−1^ is basic reproduction number [26]. Regarding the above linearized infection subsystem T and V is given by

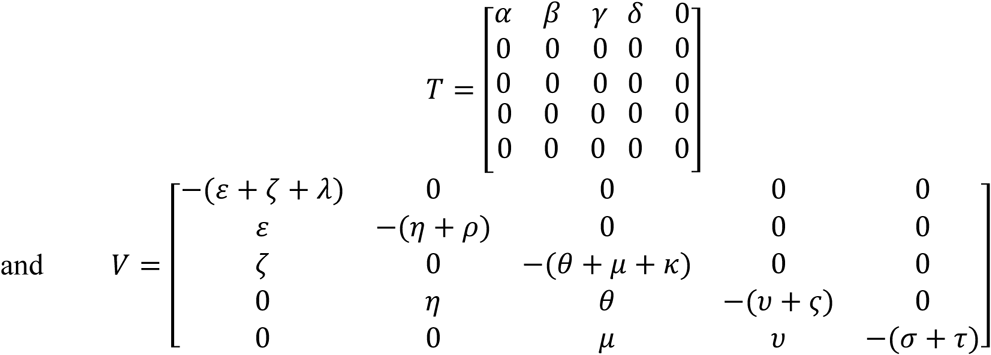

Eigenvalues of the matrix, K = −*TV*^−1^ is given by

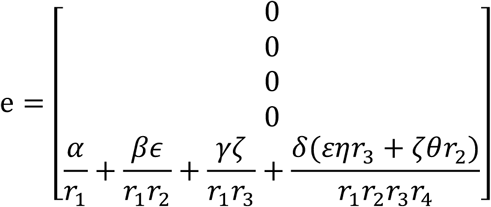

where 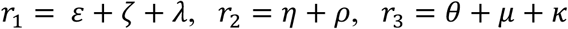 and 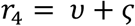

The basic reproduction number R_0_ is given by the dominant eigenvalue, hence R_0_ is defined as:

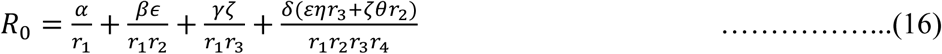

## Results

The outbreak of COVID-19 in India has been considered for the numerical simulations. The model parameters have been estimated based on data about the evolution of the pandemic in India in the period from 20 March 2020 to 31 July 2020. The impact of different countermeasures to prevent the spread has been discussed, based on the model calibration. Numerical simulations have been performed to compare the predictions of mathematical models with the real data. MATLAB 2019b has been used to simulate and analyse the results. For the study, the timeline of the various steps implemented by the GoI has been divided into seven parts i.e. phases. The first phase comprises of the time from March 20^th^, to March 24^th^, i.e. right before the implementation of the first nationwide lockdown announced by the GoI for 21 days from March 24^th^. This is taken as the Second Phase. This in turn is followed by the next, third phase, viz. the lockdown implemented from April 15^th^ to May 3^rd^, with certain relaxations from April 20^th^. The lockdown was extended yet again on May 1^st^, by the order of GoI, and the Fourth phase of our study shall comprise of the time duration from May 4^th^ to May 17^th^. May 18^th^ to May 31^st^, marks the final phase of lockdown which shall be considered as the Fifth phase. Following these, the process of un-lockdown started in the country, from June 1^st^ to June 30^th^, which shall comprise the sixth phase of our study. And the final phase i.e. Un-lockdown 2, began on July 2^nd^ till July 31^st^ with night curfews and lockdowns limited to containment zones only. This shall be our Seventh Phase [5].

The initial point of our simulation is 20 March 2020 (day 0), when India had already 195 confirmed cases reported. The initial conditions for the fraction of population in each stage is fixed as: I = 11000/population, A = 419/population, S_u_ = 10/population, D = 50/population, C = 0, R = 0, M = 0, S = 1 –I –A –S_u_ –D –C –R –M. The parameters are set as: ζ = 0.30, η = 0.189, µ = 0.0181, ν = 0.0284, τ = 0.0035, λ = 0.03, ρ = 0.03, κ = 0.0160, ς = 0.0120, σ = 0.0160. Values of the parameters for different phases are given in **Table 2**.

**Table 2:** Values of the parameters for corresponding phases.

| Phase | $\alpha$ | $\beta$ | $\gamma$ | $\delta$ | $\varepsilon$ | $\theta$ |
| --- | --- | --- | --- | --- | --- | --- |
| 1 | 0.5077 | 0.0114 | 0.407 | 0.0114 | 0.15 | 0.35 |
| 2 | 0.258 | 0.0057 | 0.15 | 0.0057 | 0.155 | 0.35 |
| 3 | 0.288 | 0.0057 | 0.186 | 0.0057 | 0.16 | 0.36 |
| 4 | 0.318 | 0.0057 | 0.218 | 0.0057 | 0.165 | 0.36 |
| 5 | 0.34 | 0.0057 | 0.24 | 0.0057 | 0.195 | 0.38 |
| 6 | 0.38 | 0.0057 | 0.28 | 0.0057 | 0.21 | 0.39 |
| 7 | 0.4 | 0.0057 | 0.3 | 0.0057 | 0.22 | 0.41 |

The basic reproduction number for Phase1 is R_0_ = 1.9730, which leads to a significant growth in the number of infected individuals.

In Phase 2, (after day 4), due to strict nationwide lockdown, basic social distancing and GoI recommendations (washing hands often, not touching one’s face, avoiding local gathering and keeping distance) the transmission of the disease was significantly reduced. The basic reproduction number is R_1_ = 0.9003 for Phase 2.

In Phase 3, (after day 25), GoI announced another nationwide lockdown with some relaxations from 20 April, which increased the disease transmission. Testing capacity of the country was also increased. Resulting basic reproduction number is R_2_ = 0.9958 for this phase.

In Phase 4, (after day 43), GoI extended the lockdown with some additional relaxations. The country has been split into three zones: red, orange and green zones. Various activities were allowed depending on the zone. GoI allowed stranded people to return their hometowns, which increased the transmission significantly. Testing capacity was also increased in this phase. The basic reproduction number for this phase is R_3_ = 1.0834.

In Phase 5, (after day 57), there was another lockdown announced by GoI with some more relaxations. In this phase red zones were further divided into containment and buffer zones. Depending upon zones, various activities were allowed. Due to plasma therapy and other therapeutic medicine, recovery rate was also improved. Basic reproduction number for phase5 is R_4_ = 1.0727.

In Phase 6, (after day 71), GoI announced reopening of the country in a phased manner with economic focus. Lockdown restrictions were only imposed in containment zones, while activities were allowed in other zones in limited mode, with night curfews (shopping malls, religious places and hotels were allowed to reopen from June 8). Basic reproduction number for phase5 is R_5_ = 1.1340.

In Phase 7, (after day 101), the second phase of the reopening began in the country. Lockdown measures were only imposed in containment zones. In all other areas most of the activities were allowed with night curfews. Basic reproduction number for phase 7 is R_6_ = 1.1431 [5].

**Figure 2A**, depicts the pandemic evolution predicted by the model with the estimated parameters up to 140 days. Solid lines represent the actual evolution of the pandemic (all cases of infection, both diagnosed and non-diagnosed cases, although the non-diagnosed cases are not counted in the data) and dashed lines represent the diagnosed pandemic evolution (all the cases that have been diagnosed and thus are reported in data). **Figure 2A** shows that in the earliest pandemic phase, the number of infected cases was underestimated. In the seventh phase, approximately 20 percent of the total cases were undetected. **Figure 2B** predicts the infected cases categorized into: non-diagnosed asymptomatic (ND AS), diagnosed asymptomatic (D AS), non-diagnosed symptomatic (ND S), diagnosed symptomatic (D S) and diagnosed with terminally ill (D IC). **Figure 2C** shows the long-term pandemic evolution over a 350-day horizon. This figure predicts that 23 percent of the population will contract the virus till 350^th^ day. The number of infected people will peak around 325^th^ day, while the peak of diagnosed cases will occur around 340^th^ day. **Figure 2D** represents long-term evolution of the infected subpopulation reaching its peak value. The comparison between the data and the prediction from the mathematical model is reported in **Figures 3**. The model predictions have been fitted on the data from 20 March 2020 to 31 July 2020 (till phase 7). **Figure 3A** shows the comparison for model results and official data for cumulative infected cases. There is some difference in model curve (blue line) and official data, due to the inconsistent reporting of cases in the official data. **Figures 3B** and **3C** represent the comparison between official data and model predictions for recovered cases and for mortal cases respectively.

**Figure 2:**
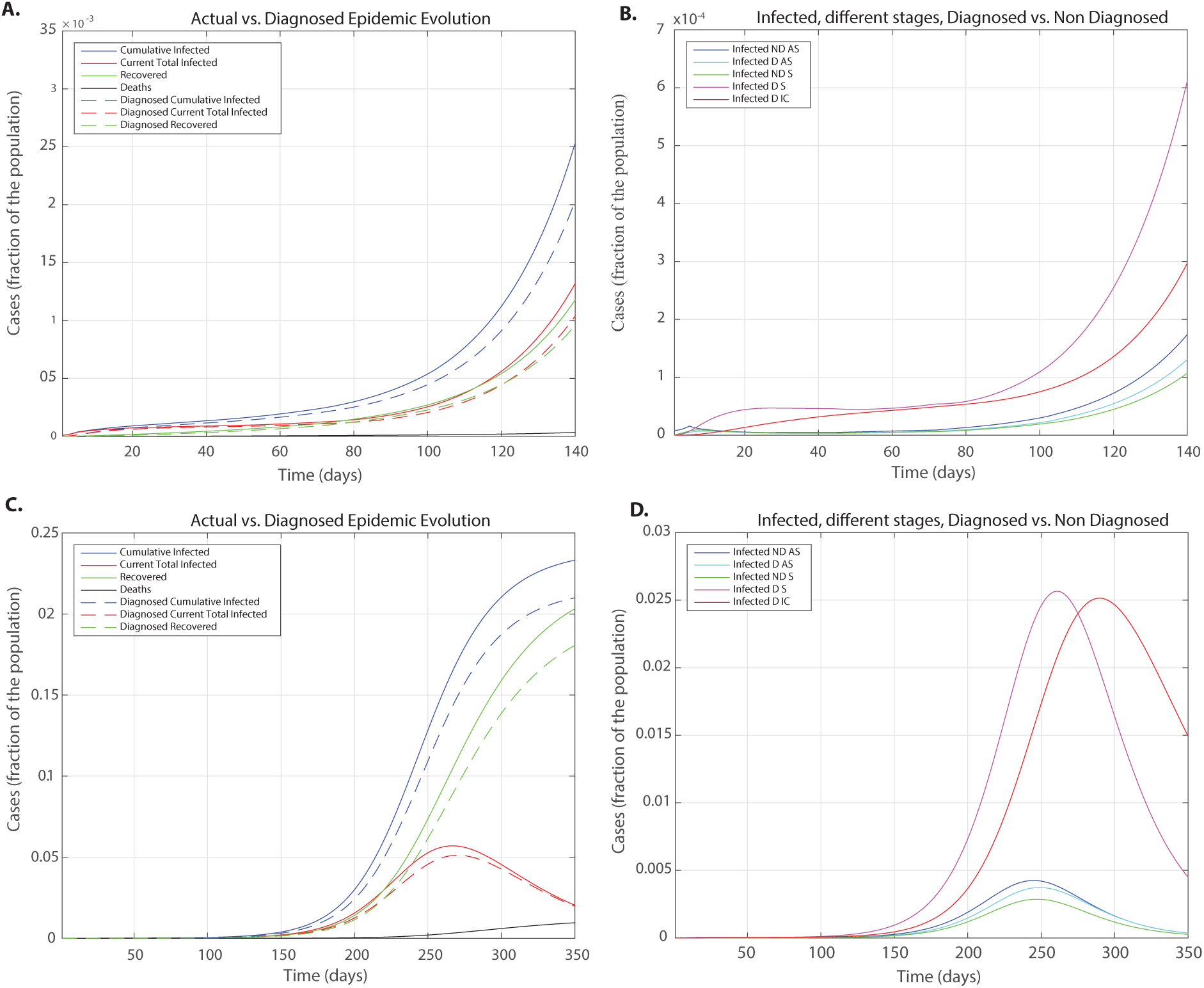
The pandemic evolution predicted by the mathematical model for the COVID-19 outbreak in India. **A:** the short-term pandemic evolution obtained by estimating the parameters based on the comparison of real time data with the model results for diagnosed cases. **B:** the different infected subpopulations for short-term horizon **C:** the long-term predicted evolution of the pandemic. **D:** the curves distinguish between different infected subpopulations for long-term horizon. The solid lines represent the actual evolution of the pandemic (this refers to all cases, both detected and undetected) and dashed lines represent the diagnosed pandemic evolution (this refers to all cases that have been detected and reported in the data).

**Figure 3:**
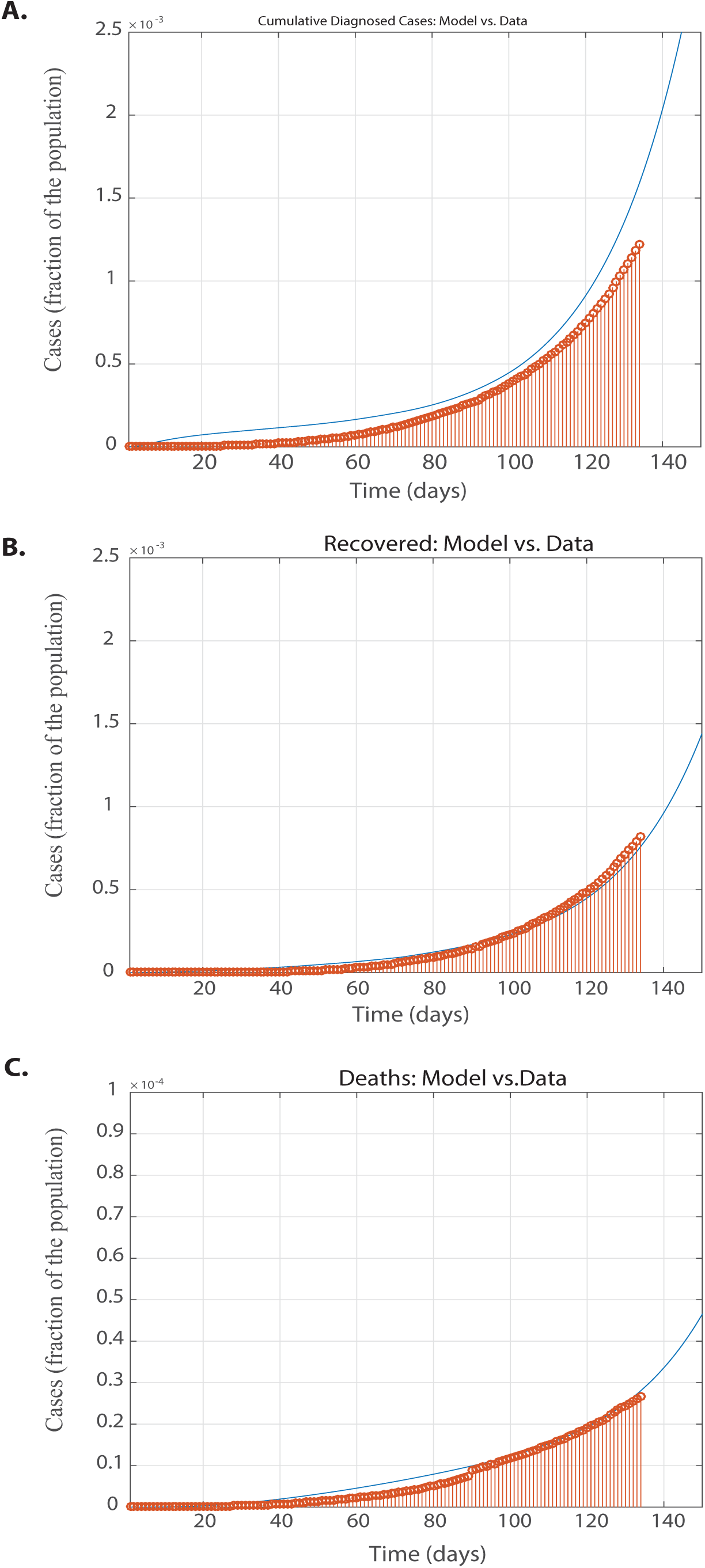
The comparison between model predictions and real time data. Red dot histograms represent the real time data and blue lines represent the model prediction. **A**: total number of reported infected cases. **B**: total number of recovered cases. **C:** total number of deaths.

If there were only social distancing countermeasures taken without any nationwide lockdowns, the model predicts that 50 percent of the population could have contracted the virus till 150^th^ day. **Figure 4** represents the evolution of pandemic without any lockdown with social distancing measures only. **Figures 4A** and **4C** show the short term and long-term pandemic evolution with social distancing measures only, respectively. **Figure 4B** and **4D** predicts the short term and long-term evolution of pandemic for infected subpopulations with social distancing measures only, respectively.

**Figure 4:**
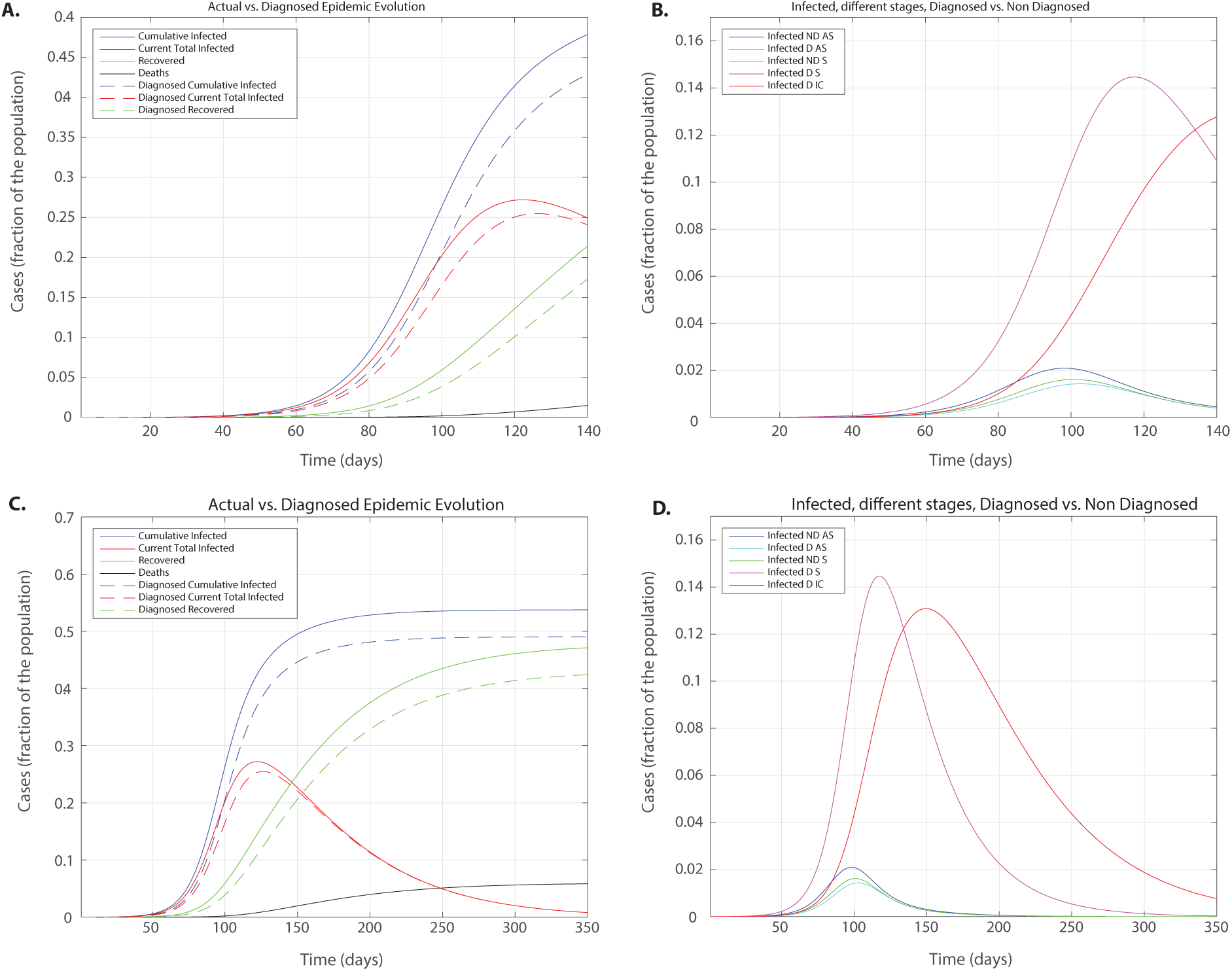
Pandemic evolution predicted by the mathematical model with social distancing measures only. **A:** the short-term predicted pandemic evolution. **B**: The curves distinguish between different infected subpopulations for the short-term predicted evolution of the pandemic **C**: the long-term predicted evolution of the pandemic **D:** the curves distinguish between different infected subpopulations for the long-term predicted evolution of the pandemic.

## Discussion

In the present study, we have considered a generalized SIDHARTHE model to quantify the effects of different lockdowns and un-lockdowns on the time evolution of COVID-19 pandemic in India. This model incorporates both diagnosed and non-diagnosed cases. The parameters were estimated by fitting the model predictions to the official data.

For the phase1, the transmission of the diseases was very high in the absence of lockdown and social distancing measures. But in the phase2, there was strict nationwide lockdown, to curb the spread of COVID-19 and reproduction number (R_1_) significantly decreased from 1.9370 to 0.9003. **Figure 2A** shows that until 25^th^ day there was very moderate increase in the curve of cumulative infected cases. From phase3 to phase5 reproduction number gradually increased from 0.9958 to 1.0727. **Figure 2A** shows that infected cases also increased gradually from 25^th^ day to 71^st^ day. In this timeline the recovery rate of the country also improved and curve representing recovery rate also started rising. With the reopening of country in phase6 and phase7, number of infected cases increased rapidly. **Figure 2A** shows that curve of cumulative infected cases increased sharply in this time. **Figure 2B** and **2D** are delineating the short-term and long-term evolution of infected subpopulations respectively with their peaks.

The distinction between diagnosed and non-diagnosed cases allows us to reckon the deviation between actual and reported diseases statistics (i.e. total infected individuals, total recovered cases, total mortality and the transmission rate). The discrepancy between actual infected (diagnosed and non-diagnosed) and official data of infected (diagnosed only) can be quantified based on this model, There were some misperceptions in the early phase of pandemic due to lack of information (many asymptomatic cases were not diagnosed), which leads to the underestimation of infected cases. Modelling results quantify the approximate error in estimating the actual number of infected cases due to inadequate number of tests being performed. The model predictions help us to explain the long-term effects of insufficient number of diagnostic tests, with sudden outbreak after a long silent period. **Figure 2C** shows the exact similar scenario of insufficient number of diagnostic tests, after the 200^th^ day there is a sudden outbreak in cases in India.

Once the parameters were estimated by fitting the model results to the clinical data, long-term pandemic evolution can be reproduced. The modelling results help us to assess and predict the effects of implementation of different strategies and protocols (i.e. first four nationwide lockdowns, extensive screening of disease, un-lockdowns). Social distancing measures are modelled by reducing the infection coefficients. Our simulated results show that different strategies implemented by GoI, have significantly delayed the peak of infected individual subpopulations. In **Figure 4C**, the peak of cumulative infected cases occurs very early in comparison to the **Figure 2C. Figure 2D** shows that nationwide lockdowns have also delayed the peak of critically ill infected people.

## Conclusion

In this paper, we have used SIDHARTHE model to discuss the potential effects of different strategies implemented by GoI on the pandemic evolution. Both, detected and un-detected cases are taken into the account. Main outcomes of the of the performed study are as follows: The basic reproduction number R_0_ decreased significantly for the second phase due to strict lockdown, but started increasing in the upcoming phases due to the various relaxations. The peaks for different infected subpopulations are considerably delayed. Underdiagnosis of the infected asymptomatic subpopulation will lead to sudden outbreak of cases in India. Without lockdown the peak of cumulative infected cases could have occurred very soon with drastic mortality rate. Our modelling results can be used for constructing efficient strategies to prevent the further spread of coronavirus in India. Our study also points out that policies such as social distancing and quarantine of the exposed population alone are not sufficient enough to end the COVID-19 pandemic in India and its states. Other stringent policies like complete lockdown as well as more testing of susceptible populations should be considered and must be incorporated systematically in mathematical models.

## Data Availability

Data has been obtained from WHO website for the considered timeline.

https://www.who.int/gho/countries/ind/data/en/

## Acknowledgement

The authors are thankful to Dr. Purnima Chaturvedi and Dr. Giulia Giordano for their valuable suggestions. Mr. Rohit Kumar acknowledges, Lakshya Avi Kishor for his help in data curation and proof reading.

## Author Contributions

Rohit Kumar: Conceptualization, data curation, methodology, simulations, preparation of result figures, writing-original draft, writing-review and editing the manuscript.

Md. Zubbair Malik: Formatting the result figures, writing-review and editing the manuscript., Sapna Ratan Shah: Supervision, writing-review and editing the manuscript.

